# Evaluation of the diagnostic value of YiDiXie™-SS, YiDiXie™-HS and YiDiXie™-D in liver cancer

**DOI:** 10.1101/2024.07.15.24310471

**Authors:** Pengwu Zhang, Huimei Zhou, Siwei Chen, Chen Sun, Zhenjian Ge, Wenkang Chen, Yingqi Li, Shengjie Lin, Wuping Wang, Yutong Wu, Xutai Li, Wei Li, Xiaoye Sun, Jinying Li, Yongqing Lai

## Abstract

**Background:** Liver cancer is one of the cancers that consistently ranks among the top five cancers in terms of incidence and mortality in many countries. However, false-positive results on enhanced CT can lead to misdiagnosis and incorrect surgery or treatment, while false-negative results on enhanced CT can lead to missed diagnosis and delayed treatment. There is an urgent need to find convenient, cost-effective and non-invasive diagnostic methods to reduce the false-positive rate of ultrasound and the false-negative and false-positive rates of enhanced CT for liver tumors. The purpose of this study is to evaluate the diagnostic value of YiDiXie™-HS, YiDiXie™-SS and YiDiXie™-D in liver tumors.

**Patients and methods:** This study finally included 217 subjects (the malignant group, n=185; the benign group, n=32). Remaining serum samples from the subjects were collected and tested using the YiDiXie™ all-cancer detection kit, which was applied to assess the sensitivity and specificity of YiDiXie™-SS, YiDiXie™-HS and YiDiXie™-D, respectively.

**Results:** The sensitivity of YiDiXie™-SS was 98.9% (96.1% - 99.8%) and its specificity was 68.8% (51.4% - 82.0%). This means that YiDiXie™-SS has very high sensitivity and high specificity in liver tumors.YiDiXie™-HS has a sensitivity of 88.1% (82.7% - 92.0%) and its specificity is 84.4% (68.2% - 93.1%). This means that YiDiXie™-HS has high sensitivity and high specificity in liver tumors.YiDiXie™-D has a sensitivity of 72.4% (65.6% - 78.4%) and its specificity is 93.8% (79.9% - 98.9%). This means that YiDiXie™-D has high sensitivity and very high specificity in liver tumors.YiDiXie™-SS has a sensitivity of 99.2% (95.8% - 100%) and a specificity of 66.7% (39.1% - 86.2%) in patients with positive enhanced CT. This means that the application of YiDiXie™-SS reduces the false-positive rate of enhanced CT by 66.7% (39.1% - 86.2%) with essentially no increase in the leakage of malignant tumors.YiDiXie™-HS has a sensitivity of 89.1% (95% CI: 78.2% - 94.9%) in patients with a negative enhanced CT, and its specificity is 85.0% (64.0% - 94.8%). This means that YiDiXie™-HS reduces the false-negative rate of enhanced CT by 85.0% (64.0% - 94.8%). YiDiXie™-D has a sensitivity of 73.1% (64.9% - 80.0%) and a specificity of 91.7% (64.6% - 99.6%) in patients with positive enhanced CT. This means that YiDiXie™-D reduces the false-positive rate of enhanced CT by 91.7% (64.6% - 99.6%). YiDiXie™-D has a sensitivity of 70.9% (95% CI: 57.9% - 81.2%) and a specificity of 95.0% (95% CI: 76.4% - 99.7%) in patients with negative enhanced CT. This means that YiDiXie ™ -D reduces the false-negative rate of enhanced CT by 70.9% (95% CI: 57.9% - 81.2%) while maintaining high specificity.

**Conclusion:** YiDiXie™-SS has very high sensitivity and high specificity in liver tumors. YiDiXie™-HS has high sensitivity and high specificity in liver tumors. YiDiXie™-D has high sensitivity and very high specificity in liver tumors. YiDiXie™-SS significantly reduces the false positive rate of liver-enhanced CT with essentially no increase in delayed treatment of malignant tumors. YiDiXie™-HS significantly reduces the false-negative rate of enhanced CT. YiDiXie™-D can significantly reduce the false-positive rate of enhanced CT or significantly reduce the false-negative rate of enhanced CT while maintaining a high specificity. The YiDiXie™ test has significant diagnostic value in liver tumors, and is expected to solve the problems of “high false-positive rate” and “high false-negative rate” of enhanced CT in liver tumors.

**Clinical trial number:** ChiCTR2200066840.

## INTRODUCTION

Hepatocellular carcinoma occurs frequently worldwide and poses a serious threat to human health and life^1^. Liver cancer is the third leading cause of cancer deaths worldwide^2,3^. The latest data show that there will be an estimated 865,000 new cases and 757,948 deaths in 2022^3^. China accounts for 45.3% of new cases and 47.1% of deaths from liver cancer worldwide^4,5^. The average number of years of life lost due to liver cancer is estimated to be 7.9 years^6^. The 5-year survival rate for advanced liver cancer is less than 20%, and the 5-year survival rate for early-stage liver cancer that undergoes radical treatment can be as high as 40%^7,8^. Some studies have shown that liver cancer is one of the most expensive cancers for men, costing as much as US$2,020.3 million in Korea alone in 2015^9^; while in countries such as the United States, the average total hospitalization cost per person is as much as US$25,738^10^. Thus, liver cancer is a serious threat to human health and brings a heavy economic burden.

Enhanced CT is widely used in the diagnosis of liver tumors. On the one hand, enhanced CT can produce a large number of false-positive results. A prospective multicenter study showed that the false-positive rate of enhanced CT in 10-20 mm liver tumors was about 23.2%^11^. When enhancement CT is positive, patients are usually treated surgically^12-14^. A false-positive result on enhanced CT means that a benign disease is misdiagnosed as a malignant tumor, and the patient will have to bear the undesirable consequences of unnecessary mental suffering, expensive surgeries and examinations, physical injuries, and even loss of function. Therefore, there is an urgent need to find a convenient, cost-effective, and noninvasive diagnostic method to reduce the false-positive rate of enhanced CT for liver tumors.

On the other hand, enhanced CT can produce a large number of false-negative results. The false-negative rate of enhanced CT is 28.4% in 20-30 mm liver tumors and 32.1% in 10-20 mm lesions^11^. When enhancement CT is negative, patients are usually taken for observation and regular follow-up^12,13^. False-negative enhancement CT results imply misdiagnosis of malignant tumors as benign disease, which will potentially lead to delayed treatment, progression of malignancy, and possibly even development of advanced stages. Patients will thus have to bear the adverse consequences of poor prognosis, high treatment costs, poor quality of life, and short survival. Therefore, there is an urgent need to find a convenient, economical and noninvasive diagnostic method to reduce the false-negative rate of enhanced CT for liver tumors.

In addition, there are some special patients who need to be extra cautious in choosing whether or not to operate, such as: smaller tumors, tumors with difficulty in preserving the liver lobe, tumors requiring lobectomy, giant liver tumors, liver insufficiency, and poor general condition of the patient. The risk of wrong surgery in these particular patients is much higher than the risk of missed diagnosis. And the false-positive result of enhanced CT means that benign diseases are misdiagnosed as malignant tumors, which will lead to misdiagnosis and wrong surgery. Therefore, there is an urgent need to find a convenient, cost-effective and noninvasive diagnostic method with high specificity to substantially reduce the false-positive rate of liver-enhanced CT in these special patients or to significantly reduce its false-negative rate while maintaining high specificity Based on the detection of miRNAs in serum, Shenzhen KeRuiDa Health Technology Co., Ltd. has developed “YiDiXie ™ all-cancer test” (hereinafter referred to as the YiDiXie ™ test)^15^. With only 200 milliliters of whole blood or 100 milliliters of serum, the test can detect multiple cancer types, enabling detection of cancer at home^15^. The YiDiXie ™ test consists of three independent tests: YiDiXie ™-HS, YiDiXie™-SS and YiDiXie™-D^15^.

The purpose of this study is to evaluate the diagnostic value of YiDiXie™-SS, YiDiXie™-HS and YiDiXie™-D in liver cancer.

## PATIENTS AND METHODS

### Study design

This work is part of the sub-study “Evaluating the diagnostic value of the YiDiXie ™ test in multiple tumors” of the SZ-PILOT study (ChiCTR2200066840).

The SZ-PILOT study (ChiCTR2200066840) is an observational, prospective, single-center research. Those who completed an informed consent form at the time of admission or physical examination for the donation of residual samples were all subjects in the study. For this investigation, the remaining 0.5 mL of serum sample was gathered.

It was a blinded trial research. The clinical information about the individuals was unknown to the laboratory staff who conducted the YiDiXie ™ test and the KeRuiDa laboratory technicians who calculated the results. YiDiXie ™ test results were likewise unknown to the clinical specialists who evaluated the participants’ clinical data.

The study was approved by the Ethics Committee of Peking University Shenzhen Hospital and was conducted in accordance with the International Conference on Harmonization for “Good clinical practice guidelines” and the Declaration of Helsinki.

### Participants

This study included participants who had a positive ultrasonography examination for liver tumors. The two groups of subjects were enrolled independently, and each subject who satisfied the inclusion criteria was added one after the other.

Inpatients with “suspected (solid or haematological) malignancy” who had provided general informed consent for the donation of the remaining samples were initially included in the study. The study classified subjects into two groups based on their postoperative pathological diagnostic: those with a diagnosis of “malignant tumor” and those with a diagnosis of “benign tumor” The study omitted subjects whose pathology findings were not entirely clear. Some of samples in the malignant group were used in our prior works^15^.

This study did not include those who failed the serum sample quality test before the YiDiXie™ test. For information on enrollment and exclusion, please see the subject group’s prior article^15^.

### Sample collection, processing

The serum samples used in this study were obtained from serum remaining after consultation and treatment, without further blood donation. Approximately 0.5 ml samples of the remaining serum from the subjects were collected in the clinical laboratory and stored at -80°C for later use in the YiDiXie™ test.

### The YiDiXie™ test

The YiDiXie ™ test is performed using the YiDiXie ™ all-cancer detection kit. The YiDiXie ™ all-cancer detection kit is developed and crafted by Shenzhen KeRuiDa Health Technology Co. It uses fluorescent PCR technology as its in-vitro diagnostic system. To determine whether cancer is present in the subject, it measures the expression levels of dozens of miRNA biomarkers in the serum.

The predefinition of appropriate thresholds for each miRNA biomarker ensures high specificity for each miRNA marker. The integration of independent assays into a parallel trial format results in a significant increase in sensitivity and high specificity for broad-spectrum cancers.

The YiDiXie ™ test consists of three distinctly different tests: YiDiXie™-Highly Sensitive (YiDiXie™ -HS), YiDiXie ™-Super Sensitive(YiDiXie ™ -SS) and YiDiXie™-Diagnosis (YiDiXie™-D). YiDiXie™-HS has been developed with sensitivity and specificity in mind. YiDiXie ™ -SS significantly increased the number of miRNA tests to achieve extremely high sensitivity for all clinical stages of all malignancy types. YiDiXie ™ -D dramatically increases the diagnostic threshold of individual miRNA tests to achieve very high specificity (very low false diagnosis rate) for all malignancy types.

Perform the YiDiXie ™ test according to the instructions provided by the YiDiXie ™ all-cancer detection kit. The detailed procedure can be found in our prior works^15^.

The laboratory technicians at Shenzhen KeRuiDa Health Technology Co., Ltd examined the raw test results and determined that the YiDiXie™ test had either “positive” or “negative” results.

### Diagnosis of enhanced CT

“Positive” or “negative” results are determined based on the diagnostic conclusion of the enhanced CT examination. If the diagnostic conclusion is “malignant or hepatocellular carcinoma (probable or highly probable)”, the test result will be judged as “positive”. If the diagnostic conclusion is “ liver tumor”, “(multiple) liver tumors (possible or probable)” or other expressions that are positive, more certain or inclined to benign tumor, or if the diagnostic conclusion is “malignant or hepatocellular carcinoma (probable) is not ruled out”, “Hepatocellular carcinoma to be ruled out”, “Liver tumor to be differentiated from hepatocellular carcinoma”, “Further investigations recommended”, etc., the diagnostic conclusion is judged to be “negative”.

### Extraction of clinical data

The subjects’ inpatient medical records or physical examination reports were used to extract clinical, pathological, laboratory, and imaging data for this study. The AJCC staging manual (7th or 8th edition) was used by trained clinicians to complete clinical staging^18,19^.

### Statistical analyses

Descriptive statistics were reported for demographic and baseline characteristics. For categorical variables, the number and percentage of subjects in each category were calculated; For continuous variables, the total number of subjects (n), mean, standard deviation (SD) or standard error (SE), median, first quartile (Q1), third quartile (Q3), minimum, and maximum values were calculated. The Wilson (score) method was used to calculate 95% confidence intervals (CIs) for multiple indicators.

## RESULTS

### Participant disposition

This study ultimately included 217 study subjects (malignant group, n = 185; benign group, n = 32 cases). The demographic and clinical characteristics of the 166 study subjects are listed in Table 1.

**Table 1.**
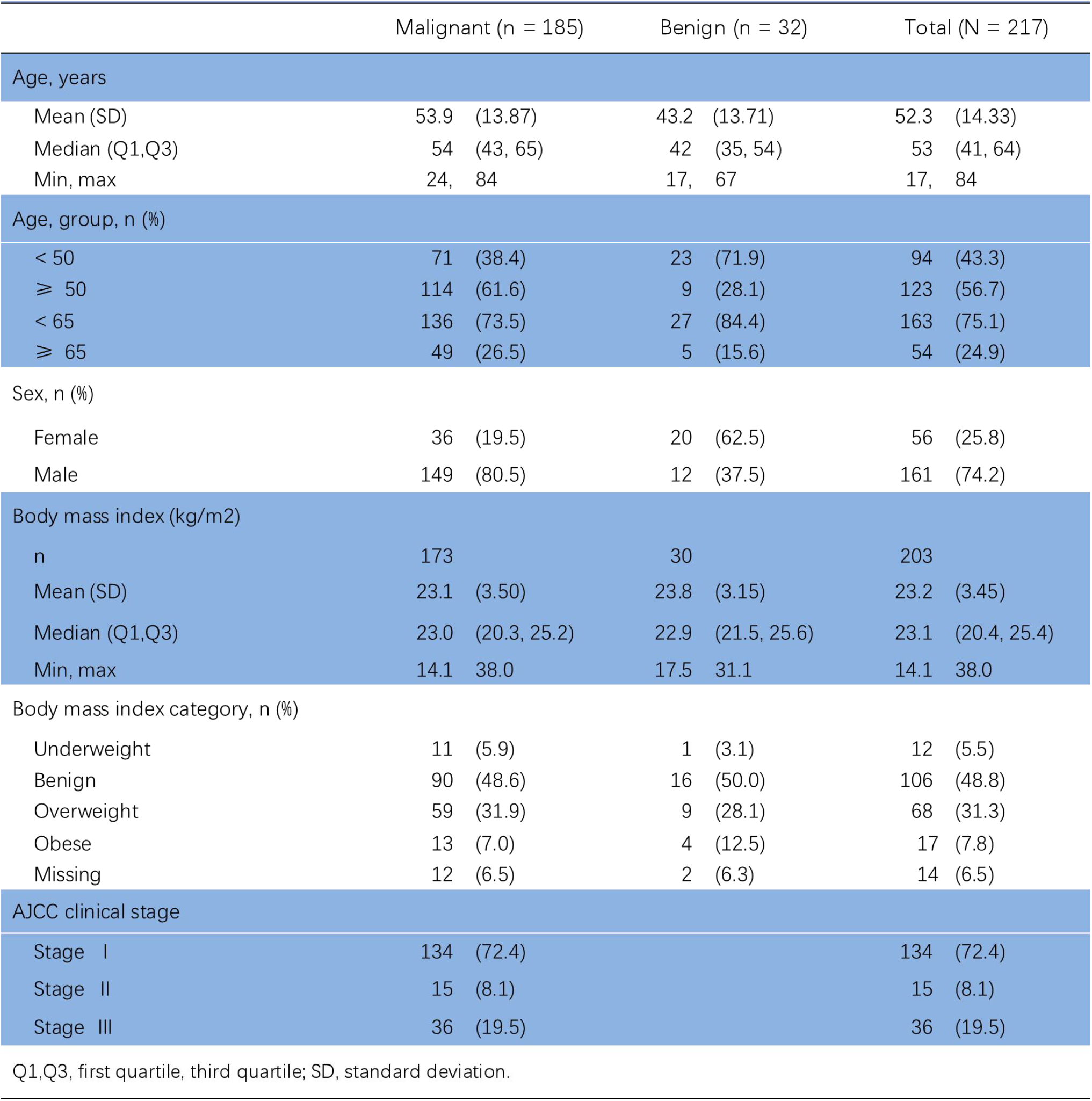
Participants’ demographic and clinical manifestation.

The two groups of study subjects were comparable in terms of demographic and clinical characteristics (Table 1). The mean (standard deviation) age was 52.3 (14.33) years and 25.8% (56/217) were female.

### Diagnostic performance of YiDiXie™-SS

As shown in Table 2, the sensitivity of YiDiXie™ -SS was 98.9% (95% CI: 96.1% - 99.8%) and its specificity was 68.8% (95% CI: 51.4% - 82.0%). This means that YiDiXie ™ -SS has very high sensitivity and high specificity in liver tumors.

**Table 2.** Performance of YiDiXie™-SS.

|  | Malignant | Benign | Total |
| --- | --- | --- | --- |
|  | 185 | 32 | 217 |
| Positive | 183 | 10 | 193 |
| Negative | 2 | 22 | 24 |

SEN, Sensitivity. SPE, Specificity. Two-sided 95% Wilson confidence intervals were calculated.

### Diagnostic performance of YiDiXie™-HS

As shown in Table 3, the sensitivity of YiDiXie™ -HS was 88.1% (95% CI: 82.7% - 92.0%) and its specificity was 84.4% (95% CI: 68.2% - 93.1%). This means that YiDiXie ™ -HS has high sensitivity and high specificity in liver tumors.

**Table 3.** Performance of YiDiXie ™ -HS.

|  | Malignant | Benign | Total |
| --- | --- | --- | --- |
|  | 185 | 32 | 217 |
| Positive | 163 | 5 | 168 |
| Negative | 22 | 27 | 49 |

SEN, Sensitivity. SPE, Specificity. Two-sided 95% Wilson confidence intervals were calculated.

### Diagnostic performance of YiDiXie™-D

As shown in Table 4, the sensitivity of YiDiXie™ -D was 72.4% (95% CI: 65.6% - 78.4%) and its specificity was 93.8% (95% CI: 79.9% - 98.9%). This means that YiDiXie ™ -D has high sensitivity and very high specificity in liver tumors.

**Table 4.** Performance of YiDiXie™-D.

|  | Malignant | Benign | Total |
| --- | --- | --- | --- |
|  | 185 | 32 | 217 |
| Positive | 134 | 2 | 136 |
| Negative | 51 | 30 | 81 |

SEN, Sensitivity. SPE, Specificity. Two-sided 95% Wilson confidence intervals were calculated.

### Diagnostic performance of hepatic enhanced CT

As shown in Table 5, the sensitivity of enhanced CT was 70.3% (95% CI: 63.3% - 76.4%;), while its specificity was 62.5% (95% CI: 45.3% - 77.1%).

**Table 5.** Performance of different Items.

| Items | Total N | Positive | Sensitivity % (95% CI)<sup>a</sup> | Total N | Negative | Specificity % (95% CI)<sup>b</sup> |
| --- | --- | --- | --- | --- | --- | --- |
| CT | 185 | 130 | 70.3% (63.3% - 76.4%) | 32 | 20 | 62.5% (45.3% - 77.1%) |
| YiDiXie™-SS in CT-positive | 130 | 129 | 99.2% (95.8% - 100%) | 12 | 8 | 66.7% (39.1% - 86.2%) |
| YiDiXie™-HS in CT-negative | 55 | 49 | 89.1% (78.2% - 94.9%) | 20 | 17 | 85.0% (64.0% - 94.8%) |
| YiDiXie™-D in CT-positive | 130 | 95 | 73.1% (64.9% - 80.0%) | 12 | 11 | 91.7% (64.6% - 99.6%) |
| YiDiXie™-D in CT-negative | 55 | 39 | 70.9% (57.9% - 81.2%) | 20 | 19 | 95.0% (76.4% - 99.7%) |

CI, confidence interval. <sup>a,b</sup> Two-sided 95% Wilson confidence intervals were calculated.

### Diagnostic performance of YiDiXie™-SS in hepatic enhanced CT-positive patients

To address the challenge of high false-positive rate of liver-enhanced CT, YiDiXie™-SS was applied to enhanced CT-positive patients.

As shown in Table 5, the sensitivity of YiDiXie™ -SS in patients with positive enhanced CT was 99.2% (95% CI: 95.8% - 100%) and its specificity was 66.7% (95% CI: 39.1% - 86.2%). This means that the application of YiDiXie ™ -SS reduces the false-positive rate of enhanced CT by 66.7% (95% CI: 39.1% - 86.2%) with essentially no increase in malignant tumor underdiagnosis.

### Diagnostic performance of YiDiXie™-HS in hepatic enhanced CT-negative patients

In order to solve the challenge of high false-negative rate of hepatic enhanced CT, YiDiXie ™ -HS was applied to hepatic enhanced CT-negative patients.

As shown in Table 5, YiDiXie ™ -HS had a sensitivity of 89.1% (95% CI: 95% CI: 78.2% - 94.9%) and its specificity was 85.0% (95% CI: 64.0% - 94.8%) in patients with negative enhanced CT. This means that the application of YiDiXie ™ -HS reduces the false negative rate of enhanced CT by 89.1% (95% CI: 95% CI: 78.2% - 94.9%).

### Diagnostic performance of YiDiXie™-D in hepatic enhanced CT-positive patients

The false-positive consequences were significantly more severe than the false-negative consequences in certain enhanced CT-positive patients, so YiDiXie ™ -D was applied to these patients to reduce their false-positive rates.

As shown in Table 5, YiDiXie ™ -D had a sensitivity of 73.1% (95% CI: 64.9% - 80.0%) and its specificity was 91.7% (95% CI: 64.6% - 99.6%) in patients with positive enhanced CT. This means that YiDiXie™-D reduces the false positive rate of enhanced CT by 91.7% (95% CI: 64.6% - 99.6%).

### Diagnostic performance of YiDiXie™-D in hepatic enhanced CT-negative patients

Certain enhancement CT-negative patients had significantly worse false-positive than false-negative consequences, so the more specific YiDiXie™-D was applied to such patients.

As shown in Table 5, YiDiXie ™ -D had a sensitivity of 70.9% (95% CI: 57.9% - 81.2%) and its specificity was 95.0% (95% CI: 76.4% - 99.7%) in patients with negative enhanced CT. This means that YiDiXie™-D reduces the false-negative rate of enhanced CT by 70.9% (95% CI: 57.9% - 81.2%) while maintaining high specificity.

## DISCUSSION

### Clinical significance of YiDiXie™-SS in hepatic enhanced CT-positive patients

The “YiDiXie ™ test” consists of 3 tests with very different characteristics: YiDiXie™-HS, YiDiXie ™-SS and YiDiXie™-D. Among them, YiDiXie™-HS combines high sensitivity and high specificity. YiDiXie ™ -SS has very high sensitivity for all malignant tumor types, but slightly lower specificity. YiDiXie ™ -D has very high specificity for all malignant tumor types, but lower sensitivity.

In patients with a positive ultrasound examination for liver tumors, the sensitivity and specificity of further diagnostic methods are important. Balancing the tension between sensitivity and specificity is essentially a balancing act between the “danger of underdiagnosis of malignant tumors” and the “danger of misdiagnosis of benign tumors”. Generally, misdiagnosis of benign liver tumors as malignant tumors usually will likely lead to unnecessary surgery, without affecting the patient’s prognosis, and their treatment costs are much lower than those of advanced cancers. In addition, the positive predictive value is higher in patients with hepatic enhanced CT positivity. It could be more harmful even if the false-negative rate is comparable to the false-positive rate. Therefore, YiDiXie ™ -SS, which has a very high sensitivity but a slightly lower specificity, was chosen to reduce the false-positive rate of hepatic enhanced CT.

As shown in Table 5, YiDiXie ™ -SS had a sensitivity of 99.2%(95% CI: 95.8% - 100%) and its specificity was 66.7%(95% CI: 39.1% - 86.2%) in patients with positive hepatic enhanced CT. The above results indicate that YiDiXie™-SS reduces the rate of hepatic enhanced CT false positives by 66.7%(95% CI: 39.1% - 86.2%) while maintaining a sensitivity close to 100%.

The above results imply that YiDiXie ™ -SS significantly reduces the probability of erroneous surgery for benign liver tumors, with essentially no increase in malignant tumor underdiagnosis. In other words, YiDiXie™-SS significantly reduces the mental suffering, expensive examination and surgery costs, radiological injuries, surgical injuries, and other adverse consequences for patients with false-positive enhanced CT, without essentially increasing the delay in the treatment of malignant tumors. Therefore, YiDiXie ™-SS meets the clinical needs well and has important clinical significance and wide application prospects.

### Clinical significance of YiDiXie™-HS in hepatic enhanced CT-negative patients

For patients with negative enhanced CT, the sensitivity and specificity of further diagnostic methods are important. Balancing the tension between sensitivity and specificity is essentially a balancing act between the “danger of malignant tumors being missed” and the “danger of benign tumors being misdiagnosed”. A higher false-negative rate means that more malignant tumors are under-diagnosed, leading to delayed treatment and progression of the malignant tumor, which may even develop into advanced stages. As a result, patients will have to bear the adverse consequences of poor prognosis, short survival, poor quality of life, and high treatment costs. In general, when benign liver tumors are misdiagnosed as malignant tumors, they are usually treated with surgery, which does not affect the patient’s prognosis, and their treatment costs are much lower than those of advanced cancers. Therefore, for patients with negative enhancement CT, the risk of malignant tumor misdiagnosis is higher than the risk of benign tumor misdiagnosis. Therefore, YiDiXie ™ -HS with high sensitivity and specificity was chosen to reduce the false-negative rate of enhanced CT for liver tumors.

As shown in Table 5, the sensitivity of YiDiXie™ -HS was 89.1%(95% CI: 95% CI:78.2% - 94.9%) and its specificity was 85.0%(95% CI: 64.0% - 94.8%) in patients with negative liver-enhanced CT. The above results indicated that the application of YiDiXie ™ -HS reduced the false-negative rate of enhanced CT by 89.1%(95% CI: 95% CI:78.2% - 94.9%).

These results imply that YiDiXie ™ -HS significantly reduces the misdiagnosis rate of a negative enhancement CT for malignant tumors. In other words, YiDiXie™-HS significantly reduces the poor prognosis, high treatment cost, poor quality of life, and short survival of patients with false-negative enhancement CT of liver tumors. Therefore, YiDiXie ™ -HS meets the clinical needs well and has important clinical significance and wide application prospects.

### Clinical significance of YiDiXie™-D

Liver tumors considered malignant usually receive surgical treatment. However, there are some conditions that require extra caution in choosing whether to operate or not, hence further diagnosis, e.g. smaller tumors, tumors with difficulty in preserving the hepatic lobe, tumors requiring lobectomy, giant liver tumors, liver insufficiency, and poor general condition of the patient.

In patients with liver tumors, both sensitivity and specificity of further diagnostic methods are important. Weighing the contradiction between sensitivity and specificity is essentially weighing the contradiction between “the danger of underdiagnosis of malignant tumors” and “the danger of misdiagnosis of benign tumors”. Because smaller tumors have a lower risk of tumor progression and distant metastasis, the “risk of “malignant tumor underdiagnosis” is much lower than the risk of “benign tumor misdiagnosis”. For tumors that are difficult to preserve the liver lobe, tumors that require lobectomy, and giant liver tumors, the risk of ““misdiagnosis of benign tumors”“ is much higher than the risk of ““misdiagnosis of malignant tumors”“ because a large amount of hepatic tissue needs to be removed, or the surgery is more traumatic. For patients with hepatic insufficiency, because the risk of “hepatic insufficiency or even hepatic failure after surgery” is higher, the “harm of misdiagnosis of benign tumor” is much higher than the “harm of misdiagnosis of malignant tumor”. For patients with poor general conditions, the risk of ‘‘misdiagnosis of benign tumor” is much higher than the risk of ‘‘malignant tumor” because the perioperative risk is much higher than the general condition. Therefore, for these patients, YiDiXie ™ -D, which has a very high specificity but a slightly lower sensitivity, was chosen to reduce the false-positive rate of liver-enhanced CT or to significantly reduce its false-negative rate while maintaining a high specificity.

As shown in Table 5, YiDiXie ™ -D had a sensitivity of 73.1% (95% CI: 64.9% - 80.0%) and a specificity of 91.7% (95% CI: 64.6% - 99.6%) in patients with positive enhanced CT, and a sensitivity of 70.9% (95% CI: 57.9% - 81.2%) in patients with negative enhanced CT. 81.2%), and its specificity was 95.0% (95% CI: 76.4% - 99.7%). These results suggest that YiDiXie ™ -D reduces the false-positive rate of enhanced CT by 91.7% (95% CI: 64.6% - 99.6%) or reduces the false-negative rate of enhanced CT by 70.9% (95% CI: 57.9% - 81.2%) while maintaining a high specificity.

These results mean that YiDiXie ™ -D significantly reduces the probability of incorrect surgery in these patients who require extra caution. In other words, YiDiXie ™ -D significantly reduces the risk of adverse outcomes such as surgical trauma, hepatic lobectomy, hepatic insufficiency, hepatic failure, and even death and other serious perioperative complications in these patients. Therefore, YiDiXie™-D meets the clinical needs well and has important clinical significance and wide application prospects.

### YiDiXie™ test has the potential to solve two challenges of liver tumor

Firstly, the three products of the YiDiXie™ test are clinically important in liver tumors. As mentioned earlier, YiDiXie™-SS, YiDiXie™-HS and YiDiXie ™ -D have significant diagnostic value in patients with positive or negative enhanced CT, respectively.

Secondly, the YiDiXie ™ test’s three products can greatly reduce the workload of clinicians and enable timely diagnosis and treatment of malignant tumor cases that would otherwise be delayed. On the one hand, YiDiXie ™ -SS provides significant relief from non-essential work for surgeons. Patients with liver tumors with positive enhanced CT are usually treated with surgery. The timely completion of these procedures is directly dependent on the number of surgeons. In many parts of the world, appointments are made for months or even more than a year. This inevitably delays the treatment of malignant cases among them, and thus it is not uncommon for patients with liver tumors awaiting surgery to develop malignant progression or even distant metastases. As shown in Table 5, YiDiXie ™ -SS reduced the false-positive rate of liver-enhanced CT by 66.7%(95% CI: 39.1% - 86.2%) with essentially no increase in missed liver cancer diagnosis. As a result, YiDiXie ™ -SS can greatly relieve the stress of non-essential work for surgeons, facilitating timely diagnosis and treatment of liver tumors or other diseases that would otherwise be delayed.

On the other hand, YiDiXie™-HS and YiDiXie™ -D can greatly reduce clinicians’ work pressure. When the diagnosis is difficult on enhanced CT, the patient usually requires an enhanced MRI or a puncture biopsy. The timely completion of these enhanced MRIs or liver puncture biopsies is directly dependent on the number of imaging physicians available. Appointments of several months or even more than a year are available in many parts of the world. It is also not uncommon for patients with liver tumors awaiting enhanced MRI examinations or puncture biopsies to experience malignant progression or even distant metastases. YiDiXie ™ -HS and YiDiXie™-D can replace these enhanced MRI examinations or puncture biopsies, greatly easing clinicians’ workload and facilitating timely diagnosis and treatment of other tumors that would otherwise be delayed.

Finally, the YiDiXie ™ test enables “just-in-time” diagnosis of liver tumors. On the one hand, the YiDiXie ™ test requires only a tiny amount of blood and allows patients to complete the diagnostic process non-invasively without having to leave their homes. Only 20 μl of serum is required to complete a YiDiXie ™ test, which is equivalent to approximately one drop of whole blood (one drop of whole blood is approximately 50 μ l, which yields 20-25 μ l of serum)^15^. Taking into account the pre-test sample quality assessment test and 2-3 repetitions, 0.2 ml of whole blood is sufficient for the YiDiXie ™ test^15^. The 0.2 ml of finger blood can be collected at home by the average patient using a finger blood collection needle, eliminating the need for venous blood collection by medical personnel, and allowing patients to complete the diagnostic process non-invasively without having to leave their homes^15^.

On the other hand, the diagnostic capacity of the YiDiXie ™ test is virtually unlimited. Figure 1 shows the basic flowchart of the YiDiXie ™ test, which shows that not only does the YiDiXie™ test not require a doctor or medical equipment, but it also does not require medical personnel to collect the blood.

**Figure 1.**
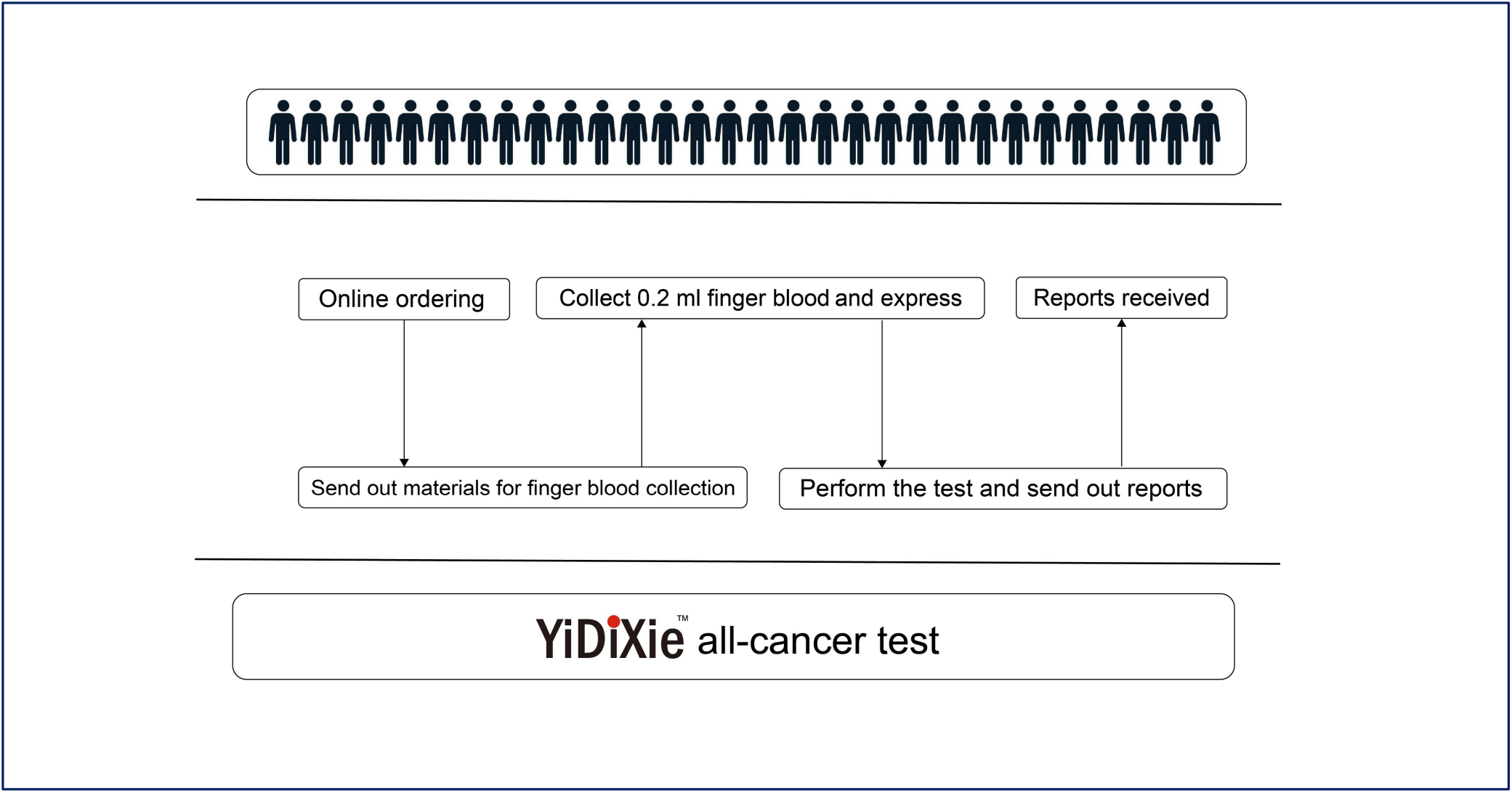
Basic flowchart of the “YiDiXie™ test”.

As a result, the YiDiXie ™ test is completely independent of the number of medical personnel and facilities, and its capacity is virtually unlimited. In this way, the YiDiXie ™ test enables “just-in-time” diagnosis of liver tumors, without patients having to wait anxiously for an appointment.

In short, the YiDiXie ™ test has significant diagnostic value in liver cancer, and is expected to solve the problems of “ high false-positive rate of hepatic enhanced CT “ and “ high false-negative rate of hepatic enhanced CT”.

### Limitations of the study

Firstly, the number of cases in this study was small, so further clinical studies with larger sample sizes are necessary for further assessment.

Secondly, the purpose of this study was to control for both malignant and benign tumors in inpatients, and future cohort studies of patients with positive thyroid tumor ultrasound scans are needed to further assess them.

Finally, the results of this study may be biased because it was a single-centre study. Future studies in multiple centers are necessary to further evaluate it.

## CONCLUSION

YiDiXie™-SS has very high sensitivity and high specificity in liver tumors. YiDiXie ™ -HS has high sensitivity and high specificity in liver tumors. YiDiXie ™ -D has high sensitivity and very high specificity in liver tumors. YiDiXie™-SS significantly reduces the false positive rate of liver-enhanced CT with essentially no increase in delayed treatment of malignant tumors. YiDiXie ™ -HS significantly reduces the false-negative rate of enhanced CT.YiDiXie ™ -D can significantly reduce the false-positive rate of enhanced CT or significantly reduce the false-negative rate of enhanced CT while maintaining a high specificity. The YiDiXie ™ test has significant diagnostic value in liver tumors, and is expected to solve the problems of “ high false-positive rate” and “high false-negative rate” of enhanced CT in liver tumors.

## Data Availability

All data produced in the present study are contained in the manuscript.

